# Nasopharyngeal aspirates and stool sample collection for molecular tuberculosis testing in children with severe pneumonia – a mixed methods study

**DOI:** 10.64898/2026.09.24.26363964

**Authors:** Bandana Bhatta, Dim Bunnet, Mikaela Coleman, Douglas Mbang Massom, Joel Djaha, Marie-France Larissa Banga, Sanary Kaing, Yara Voss de Lima, Kabuswe Mbalazi, Eric Komena, Sylvie Kwedi-Nolna, Saniata Cumbe, Naome Natukunda, Perfect Shankalala, Mastula Nanfuka, Clementine Roucher, Manon Lounnas, Minh Huyen Ton Nu Nguyet, Aurelia Vessière, Anne-Laure Maillard, Eden Ngu, Basant Joshi, Laurence Borand, Raoul Moh, Celso Khosa, Chishala Chabala, Juliet Mwanga-Amumpere, Jean-Voisin Taguebue, Eric Wobudeya, Maryline Bonnet, Olivier Marcy, Joanna Orne-Gliemann, the TB-Speed Pneumonia Study Group

## Abstract

**Background:** Child-friendly sample collection methods are needed to improve microbiological tuberculosis diagnosis in children. We assessed the feasibility, diagnostic yield, safety, tolerability, and acceptability of nasopharyngeal aspirates (NPA) and stool sample collection for XpertMTB/RIF Ultra (Ultra) testing in children <5 years with severe pneumonia in six high tuberculosis incidence countries.

**Methods:** We conducted a cross-sectional mixed-methods study (10/2020-03/2021) nested within the TB-Speed Pneumonia trial (NCT03831906) in 15 tertiary hospitals. Uptake and diagnostic yield of NPA and stool, sampled by trained nurses, were computed. Nurses documented NPA feasibility and related adverse events. NPA tolerability was assessed through discomfort/distress/pain scales completed by nurses, parents/caregivers, and children >3 years. Individual interviews investigated nurses’ and parents/caregivers’ perceptions and experiences of both procedures.

**Results:** Of 542 children enrolled, 541 (99.8%) and 423 (75.5%) had NPA and stool samples successfully collected, respectively; 8 (1.5%) had positive Ultra on any sample, 8 (1.5%) on NPA, and 3 (0.6%) on stool samples. Nurses reported that NPA was difficult to collect in 27 (5.0%) children. 130 (24%) children had bloodstained NPA samples and <5% experienced transient heart rate deceleration, digestive or respiratory distress during NPA. Median nurse-assessed tolerability scores were 1/10 [0,3] before and 5/10 [2,6] during NPA. Although most respondents perceived NPA as painful, all appreciated that NPA aimed to improve child health. Nurses reported that NPA and stool collection was quicker, less invasive than gastric aspiration. Parents/caregivers trusted nurses’ skills and valued the novelty of NPA. All respondents easily engaged in stool collection, but were frustrated to depend on children passing stool. Nurses underlined that continuous staff training and mentoring would sustain feasibility and self-efficacy in obtaining adequate samples.

**Conclusions:** NPA and stool are viable specimen collection options for microbiological diagnosis of tuberculosis in young children <5 years with severe pneumonia in high tuberculosis incidence countries.

## Introduction

There were an estimated 1.2 million cases of tuberculosis among children and young adolescents worldwide in 2024, but less than half were notified to the World Health Organisation (WHO), mostly because they were not diagnosed (1). As a result, a large majority of the 172,000 children who died from tuberculosis, were likely never offered treatment for this curable and preventable disease (1, 2). Tuberculosis diagnosis is challenging in young children, due to the difficulties for children to self-expectorate good quality respiratory samples for microbiological detection of *Mycobacterium tuberculosis,* and due to the paucibacillary nature of tuberculosis in this age group, all of which results in a low microbiological yield (3).

In 2022, WHO recommended the use of innovative and child-friendly respiratory specimen collection methods for rapid molecular testing using Xpert MTB/RIF Ultra (4). Traditional respiratory sample collection methods for young children such as induced sputum or gastric aspiration are indeed complicated to implement in routine practice. Alternative sample collection methods include nasopharyngeal aspiration (NPA) and stool collection, which aims at retrieving bacteria from the respiratory track or that have been ingested (5, 6). WHO recently recommended the combination of rapid molecular testing on one respiratory sample and on stool sample in children with presumptive tuberculosis (7, 8) .

Although tuberculosis classically presents as a sub-acute pulmonary disease, in high tuberculosis incidence countries, children with tuberculosis may also present with severe acute pneumonia (9). A meta-analysis showed that up to 23% of children under 5 years of age admitted to hospital with pneumonia are eventually diagnosed with tuberculosis (10). However, current recommendations advise assessing for tuberculosis in children only if symptoms of pneumonia persist or if antibiotic treatment is ineffective (4), which may result in missing or delaying tuberculosis treatment in highly vulnerable children (9). The TB-Speed Pneumonia study evaluated the effect on mortality of adding systematic tuberculosis detection using Ultra on one NPA and one stool sample in any child <5 years presenting with severe pneumonia in 6 high tuberculosis incidence and resource-limited countries. Although improved child survival from the intervention did not reach statistical significance, there was a trend toward mortality reduction in children with co-occurrence of severe pneumonia and severe acute malnutrition (SAM) in addition to an increase in microbiological tuberculosis confirmation (11).

The combination of NPA and stools could be a strategy for microbiological testing of children with severe pneumonia. However, NPA could be poorly tolerated or more difficult to perform in children with dyspnoea and could be poorly accepted by healthcare workers (HCWs) and families (12). Stool sample collection, although considered safe and highly feasible, could also face acceptability and implementation challenges. More evidence is needed around NPA and stool sample collection in this vulnerable group to inform policy makers, programme managers and implementers. In this study, we sought to describe the uptake of NPA and stool sample collection among children with severe pneumonia; their microbiological yield; the operational feasibility (process, logistics), reported and observed tolerability (pain/distress/discomfort) and safety of NPA in this group; and the perceived feasibility and acceptability (point of view, experience, perceptions and feelings) among HCWs and parents/caregivers of NPA and stool sample collection in children with severe pneumonia.

## Methods

### Study design, settings, and populations

We conducted a cross-sectional concurrent mixed methods study, nested in the TB-Speed Pneumonia stepped-wedge cluster-randomized trial (NCT03831906, https://clinicaltrials.gov/study/NCT03831906), that assessed the effect of adding systematic Ultra testing of NPA and stool samples on 12-week all-cause mortality in children presenting with WHO-defined severe pneumonia (11, 13). The trial was conducted in 15 tertiary hospitals within five sub-Saharan Africa countries (Cote d’Ivoire, Cameroon,

Mozambique, Uganda and Zambia) and one country in Asia (Cambodia) with high tuberculosis incidence. This nested study, conducted over the last 6 months of the trial (October 2020 to March 2021), included: i) trial data to assess sample collection uptake and diagnostic yield among children aged 2 to 59 months, hospitalized with WHO-defined severe pneumonia and enrolled within the intervention arm, ii) repeated questionnaires collected among all nurses in charge of collecting NPA and stool during the nested study period, iii) NPA pain scales completed by parents/caregivers, nurses, and children themselves when >3 years, and iv) individual interviews among nurses and parents/caregivers purposively selected among the 15 study sites based upon their diversity of socio-demographic characteristics (i.e., age, sex, educational background and employment status).

### Study procedures

At baseline visit, after parent/caregiver informed consent, inpatient children underwent a complete clinical evaluation, a digital chest X-ray, blood draws including HIV testing. Microbiological specimen collection was performed as soon as possible and within 24 hours of hospital admission. It included one NPA sample collected by the nurse on the day of admission, and one stool sample collected as soon as the child was able to produce stool. NPA was performed, without prior nasal instillation, under peripheral oxygen saturation monitoring with an oximeter by trained study or ward nurses. They used a mucus aspirator (catheter size French 8) connected to a suction device in order to stimulate the cough reflex and obtain 2-5 ml of mucus. The procedure was repeated in the second nostril if unsuccessful or with insufficient volume. Ultra testing on NPA was performed immediately, either at the hospital laboratory with the GeneXpert automate, or undertaken in the ward or in a side-laboratory next to the ward using a battery operated one-module device (G1 Edge®, Cepheid, SunValley, US). Ultra testing on stool, which requires prior processing, was performed at the hospital laboratory (13).

### Data collection

Data on the feasibility, microbiological yield and safety of NPA and stool sample collection were extracted from the TB-Speed Pneumonia trial clinical database. The safety assessment of NPA sample collection relied upon nurses completing an adverse event form for each child after sample collection and assessments of event association with the NPA/stool sample process. Nurses reported on the operational feasibility of NPA and difficulties experienced during each child’s NPA sample collection in a dedicated questionnaire completed after each NPA attempt; medical investigators reported Serious Adverse Events (SAEs) if considered life-threatening (14). Tolerability of NPA sampling was assessed before and during the procedure, as a discomfort/distress/pain score using three different scales initially developed and used for pain evaluation (**Supplementary figure 1**): i) a Visual Analogue Scale (VAS scale) used by the parents/caregivers present at the time of the procedure; ii) the Face-Leg-Activity-Cry-Consolability behavioural scale (FLACC scale; **Supplementary table 1**) (15) used by nurses; and iii) the Wong Baker Face Scale (WBF scale), allowing self-assessment of discomfort/distress/pain by children above 3 years of age (16).

Perceived feasibility and acceptability of NPA and stool sampling was explored during individual interviews among parents/caregivers and nurses. Interviews were led by experimented (Masters-level) and specifically trained social science research assistants (mixed gender across the study countries). Social science research assistants in each country liaised with study investigators and study nurses, to contact eligible nurses and parents/caregivers. Written informed consent was obtained before the interview. Interviews were conducted within the study sites, using a semi-structured guide co-designed by the principal investigators (PhD, MD) and the social science research assistants. Interviews were audio-recorded with permission of respondents. Interview audios, transcripts and translated transcripts (if interview was conducted in a language other than English) were uploaded on a secure server maintained at University of Bordeaux, France. The duration of interviews usually ranged between 1 hour and 1 hour and 30 minutes.

### Outcomes

Outcomes were i) the feasibility of NPA and stool sampling in terms of specimen collection attempt, sampling and testing uptake; ii) the operational feasibility of NPA in terms of reported technical or logistic difficulties faced during the procedure; iii) the Ultra detection yield; iv) the tolerability of NPA sampling as a 0-10 integer score measuring change in discomfort/distress/pain measure before and during the procedure; v) the safety of NPA in terms of occurrence of adverse events of interest (respiratory, cardiac, digestive events, and nose bleeding); vi) the perceived feasibility and acceptability of NPA and stool sample collection defined as the extent to which the nurses delivering or the child/parents/caregivers experiencing the specimen collection procedures considered them to be a) appropriate, based on their anticipated or experiential cognitive and emotional responses (following the Sekhon Theoretical Framework of Acceptability – TFA (17)) and b) doable in their context.

### Data analysis

#### Quantitative data analysis

Feasibility outcomes, detection yield and adverse events were described as frequency and proportions. Uptake and diagnostic yield were compared between NPA and stool sampling using Chi square or Fisher’s exact tests. Tolerability of NPA sampling was measured as absolute median (and interquartile range – IQR) scores and as difference in discomfort/distress/pain scores before and during the NPA procedure obtained for each scale. Analyses were adjusted for study site; subgroup analyses were conducted to assess the effect of hospital NPA procedural experience on NPA tolerability, using length of time in the intervention arm as the surrogate measure of experience (or expertise/proficiency). Determinants of nurse-assessed poor tolerability of NPA were identified from *p-*value estimates using non-parametric tests Wilcoxon Rank-Sum for binary categorical data and Kruskal-Wallis for multiple categorical data. Factors associated with poor tolerability were further assessed by multivariable logistic regression, after excluding four hospitals reporting outlying FLACC assessment, consistently reporting no difference or improved (decreased) discomfort/distress/pain scores during the procedure compared to before. All quantitative analyses were performed using R software (version 4.3.2).

#### Qualitative data analysis

Interview transcripts were analysed through a combination of theory-led (deductive, i.e., pre-defined perceived feasibility topics in the guide and the TFA) and data-led approaches (inductive, i.e., new themes emerging from the interviews). Two separate codebooks were developed for parents/caregivers’ and nurses’ interviews. Data was coded by social sciences research assistants using Nvivo15® qualitative data management software. Thematic summaries on the perceived feasibility and acceptability of NPA and stool sampling were prepared separately for parents/caregivers and nurses. Analysis and interpretation were conducted between social science research assistants, the trial and study coordinators and co-investigators (PhD and MD), who met during weekly online meetings.

### Patient and Public Involvement

Study findings were presented to key study stakeholders (including the facility in charge from the study sites and patient representatives) prior manuscript write-up.

### Ethical approval

The TB-Speed Pneumonia trial was approved by the WHO Ethics Review Committee (protocol ID: TB-Speed Pneumonia, international version 2.0 dated 22/11/2018), the French National Institute for Health and Medical Research (Inserm) Ethics Committee (IRB00003888) (protocol ID: C18–26), and national ethics committees and relevant regulatory authorities in each participating country.

## Results

From October 1, 2020 to March 31, 2021, a total of 542 children hospitalised with severe pneumonia of median age 11 (IQR 6,18) months, 309 (57%) male, 120 (22.1%) with SAM, and 19 (3.5%) living with HIV, were recruited in the sub-study. As per the stepped-wedge trial design, the 15 participating hospitals (**Table 1**) had been in the intervention phase for a median of 210 days (IQR 107, 341) before the nested study commenced (**Supplementary table 2**). Therefore, 229 (42.4%) children were recruited in hospitals with less than 6 months of experience of using NPA and stool sampling (“novice” hospitals).

**Table 1.** Baseline (demographic, institutional, respiratory & comorbidity) characteristics of children included in sub-study.

| Characteristics | N* if different<br>from N | N=542<br>N (%) or median [IQR] |
| --- | --- | --- |
| Sex male |  | 309 (57.0) |
| Age (months) |  | 11 [6,18] |
| <1 year |  | 288 (53.1) |
| ≥1year |  | 254 (46.9) |
| Country & Hospital |  |  |
| Cambodia |  | 68 (12.5) |
| Referral Hospital, Kampong Cham |  | 35 (6.5) |
| National Paediatric Hospital, Phnom Penh |  | 23 (4.2) |
| Takeo Referral Hospital, Takeo |  | 10 (1.8) |
| Cameroon |  | 108 (19.9) |
| Chantal Biya Foundation, Yaoundé |  | 58 (10.7) |
| District Hospital Biyem Assi, Yaoundé |  | 50 (9.2) |
| Côte d’Ivoire |  | 87 (16.1) |
| Angré UTH, Abidjan |  | 5 (0.9) |
| Treichville UTH, Abidjan |  | 39 (7.2) |
| Cocody UTH, Abidjan |  | 43 (7.9) |
| Mozambique |  | 33 (6.1) |
| Central Hospital, Maputo |  | 25 (4.6) |
| Jose Macamo General Hospital, Maputo |  | 8 (1.5) |
| Uganda |  | 159 (29.3) |
| Mulago National Referral Hospital, Kampala |  | 39 (7.2) |
| Holy Innocents Childrens' Hospital, Mbarara |  | 75 (13.9) |
| Regional Reference Hospital, Jinja |  | 45 (8.3) |
| Zambia |  | 87 (16.1) |
| University Teaching Hospital, Lusaka |  | 41 (7.6) |
| Arthur Davidson Children Hospital, Ndola |  | 46 (8.5) |
| Time from hospital allocation to the intervention arm to 1<sup>st</sup> enrolment in sub-study (days) |  | 210 [106.8, 341.0] |
| Hospital experience in performing NPA |  |  |
| Novice (<6 months experience) |  | 229 (42.4) |
| Proficient (>6 months experience) |  | 312 (57.6) |
| Respiratory rate (breaths/min) |  | 46 [38,55] |
| Tachypnea | 541 | 185(34.2) |
| Heart rate (beats/min) |  | 145 [130,160] |
| Peripheral O2 saturation (%) |  | 96 [93,98] |
| SpO2<90% | 56 | 87 [84.75,88.25] |
| Severe acute malnutrition** |  | 120 (22.1) |
| HIV positive | 533 | 19 (3.5) |

\* SAM defined according to WHO guidelines as weight for Height Z score < 3 DS or MUAC<115mm or bilateral
oedema

### Sample collection feasibility, safety and yield

Of 542 children enrolled, 541 (99.8%) and 423 (78.0%) children had NPA and stool successfully collected, respectively (p<0.0001) (**Figure 1**). Children’s behaviour was considered very easy (n=159, 29.3%) or manageable (n=352, 64.9%) during NPA collection by the nurses, respectively (**Table 2**).

**Figure 1.**
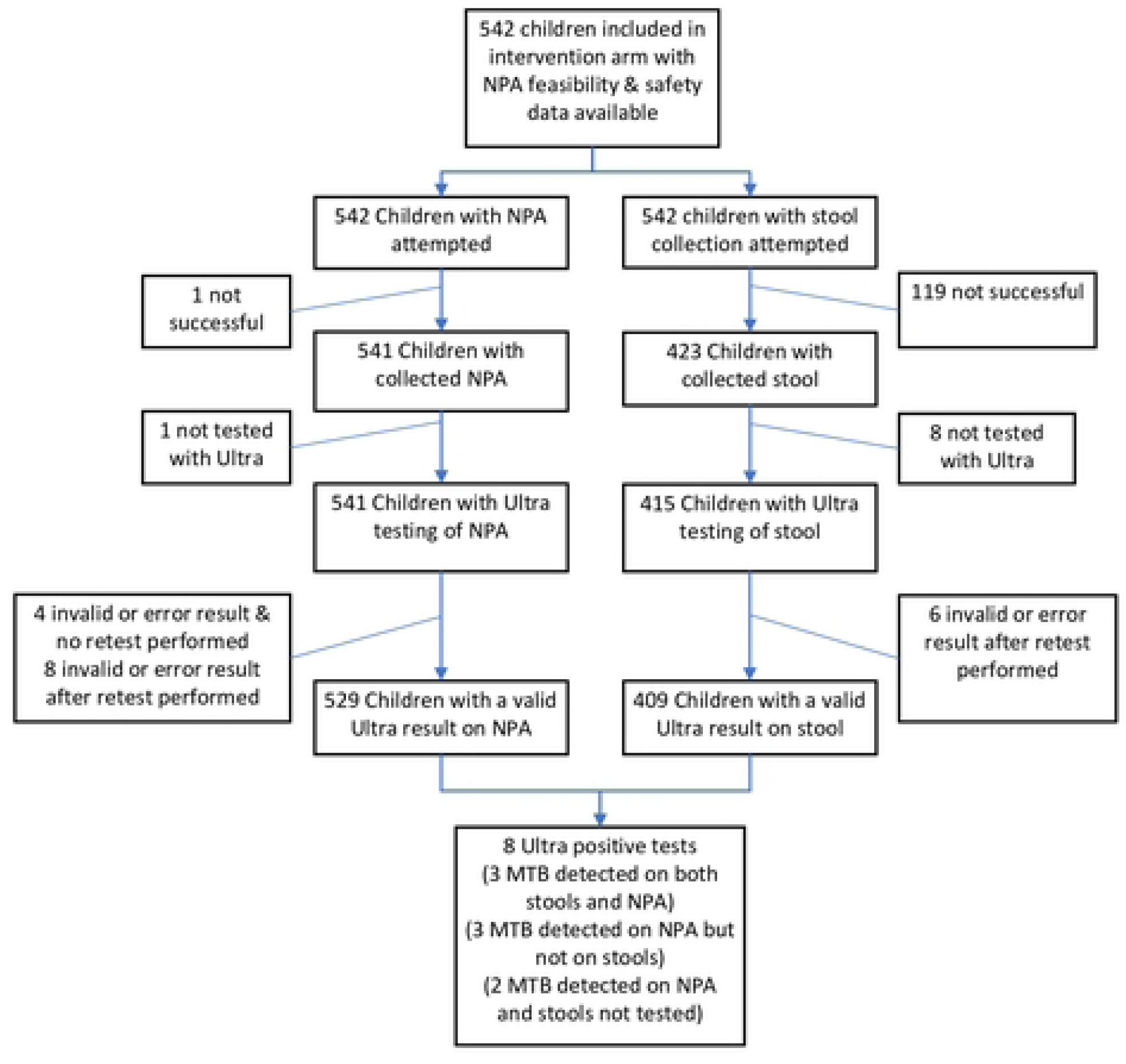
Flowchart of the feasibility and microbiological yield of Naso-Pharyngeal Aspirate (NPA) and stool sample collection

**Table 2.**
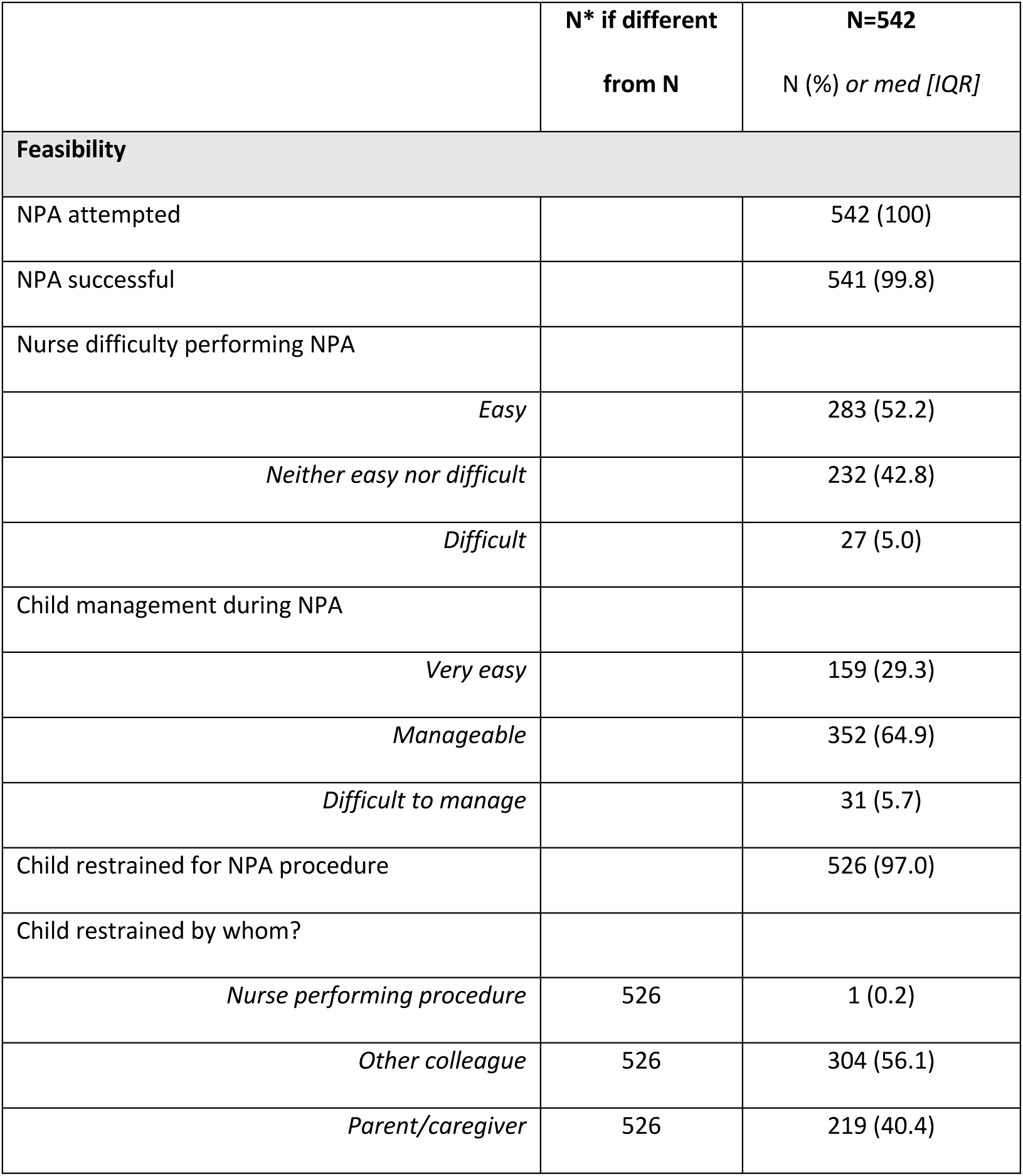
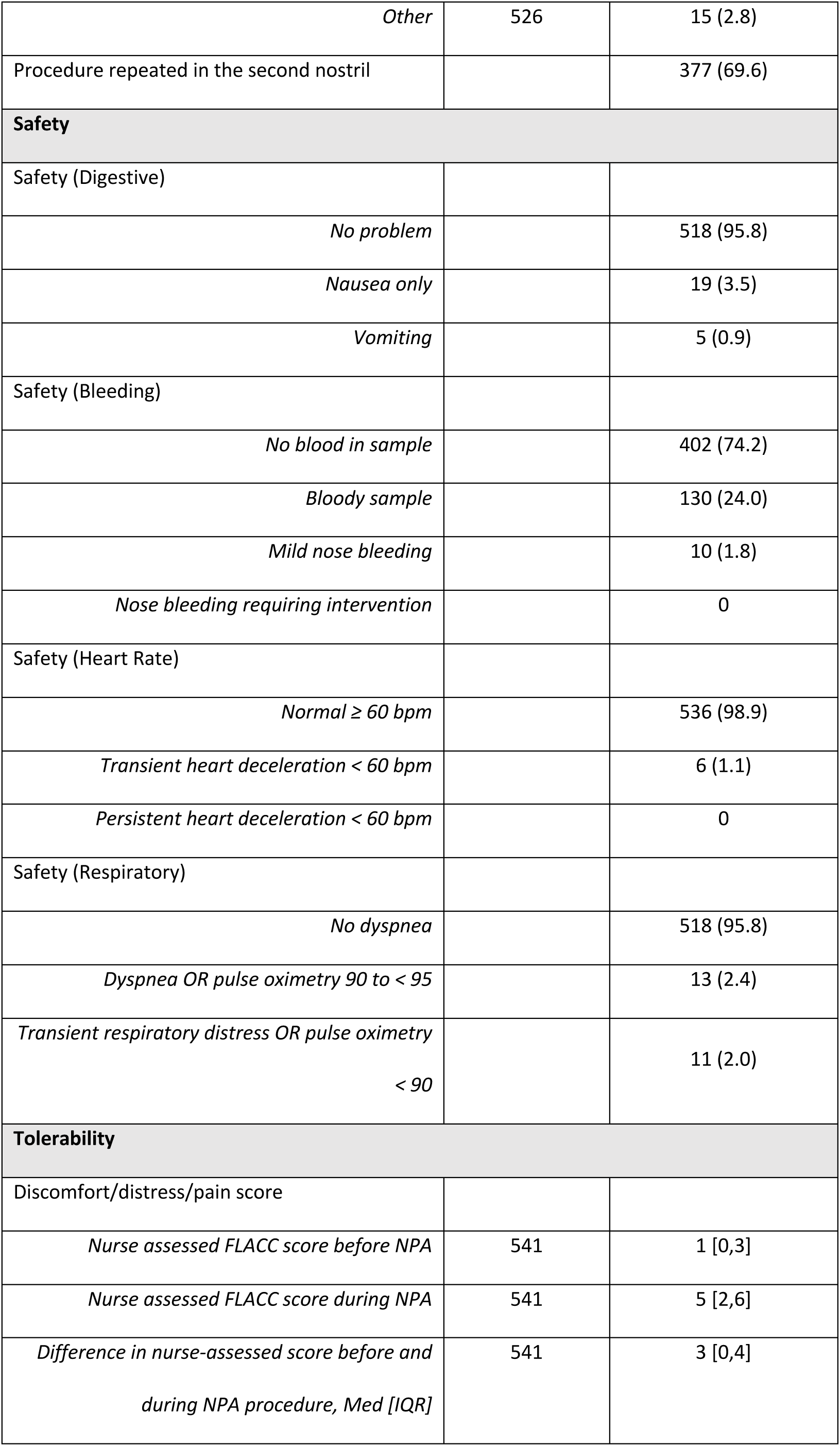
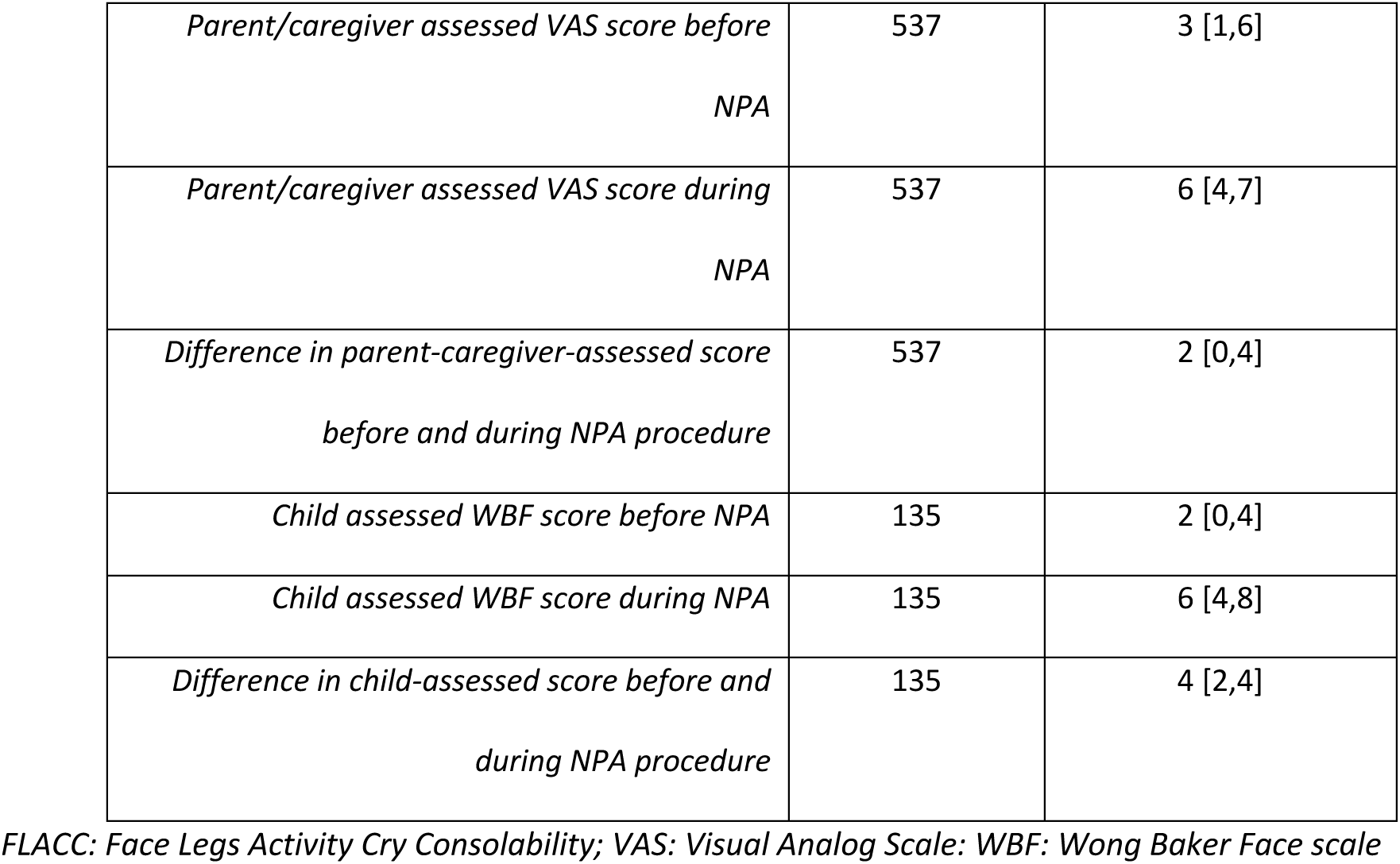
Nurse-reported feasibility and safety of Naso-pharyngeal aspirate (NPA) sample collection, and NPA tolerability scores assessed by nurses, parents/caregivers and children.

|  | N* if different<br>from N | N=542<br>N (%) or med [IQR] |
| --- | --- | --- |
| **Feasibility** |  |  |
| NPA attempted |  | 542 (100) |
| NPA successful |  | 541 (99.8) |
| Nurse difficulty performing NPA |  |  |
| *Easy* |  | 283 (52.2) |
| *Neither easy nor difficult* |  | 232 (42.8) |
| *Difficult* |  | 27 (5.0) |
| Child management during NPA |  |  |
| *Very easy* |  | 159 (29.3) |
| *Manageable* |  | 352 (64.9) |
| *Difficult to manage* |  | 31 (5.7) |
| Child restrained for NPA procedure |  | 526 (97.0) |
| Child restrained by whom? |  |  |
| *Nurse performing procedure* | 526 | 1 (0.2) |
| *Other colleague* | 526 | 304 (56.1) |
| *Parent/caregiver* | 526 | 219 (40.4) |
| *Other* | 526 | 15 (2.8) |
| Procedure repeated in the second nostril |  | 377 (69.6) |
| **Safety** |  |  |
| Safety (Digestive) |  |  |
| *No problem* |  | 518 (95.8) |
| *Nausea only* |  | 19 (3.5) |
| *Vomiting* |  | 5 (0.9) |
| Safety (Bleeding) |  |  |
| *No blood in sample* |  | 402 (74.2) |
| *Bloody sample* |  | 130 (24.0) |
| *Mild nose bleeding* |  | 10 (1.8) |
| *Nose bleeding requiring intervention* |  | 0 |
| Safety (Heart Rate) |  |  |
| *Normal $\geq 60$ bpm* |  | 536 (98.9) |
| *Transient heart deceleration $< 60$ bpm* |  | 6 (1.1) |
| *Persistent heart deceleration $< 60$ bpm* |  | 0 |
| Safety (Respiratory) |  |  |
| *No dyspnea* |  | 518 (95.8) |
| *Dyspnea OR pulse oximetry 90 to $< 95$* |  | 13 (2.4) |
| *Transient respiratory distress OR pulse oximetry $< 90$* |  | 11 (2.0) |
| **Tolerability** |  |  |
| Discomfort/distress/pain score |  |  |
| *Nurse assessed FLACC score before NPA* | 541 | 1 [0,3] |
| *Nurse assessed FLACC score during NPA* | 541 | 5 [2,6] |
| *Difference in nurse-assessed score before and during NPA procedure, Med [IQR]* | 541 | 3 [0,4] |
| *Parent/caregiver assessed VAS score before*<br>*NPA* | 537 | 3 [1,6] |
| *Parent/caregiver assessed VAS score during*<br>*NPA* | 537 | 6 [4,7] |
| *Difference in parent-caregiver-assessed score*<br>*before and during NPA procedure* | 537 | 2 [0,4] |
| *Child assessed WBF score before NPA* | 135 | 2 [0,4] |
| *Child assessed WBF score during NPA* | 135 | 6 [4,8] |
| *Difference in child-assessed score before and*<br>*during NPA procedure* | 135 | 4 [2,4] |

FLACC: Face Legs Activity Cry Consolability; VAS: Visual Analog Scale; WBF: Wong Baker Face scale

Restraining of the child was used in 526 (97%) cases, and a repeat procedure in the second nostril was required in 377 (69.6%) children to obtain sufficient sample volume. Severe adverse events (grade 3) occurred in 11 (2.4%) children sampled with NPA and were due to transient respiratory distress or oxygen saturation reduction below 90%. There were no serious adverse events. Grade 1 (mild) nausea or vomiting occurred in less than 5% of children. Of the 130 (24%) children who had blood in their NPA sample, only 10 (1.8%) had mild nose bleeding, and no bleeding management intervention was required in any case. Blood in sample was more common in children ≥1 year of age (n=76/254, 29.9%) than in younger children <1 year (n=53/288, 18.8%) (p-value=0.002) (**Supplementary table 3**).

Overall, 529 (97.6%) and 409 (75.5%) children had a valid Ultra result on NPA and stools, respectively (p<0.0001) (**Figure 1**). Of 542 children with either NPA or stool attempted, 8 (1.5%) children had positive Ultra on any sample, including 8 (1.5%) on NPA samples and 3 (0.6%) on stool samples (p=0.13). Three children had positive results on NPA only (**Figure 1**).

### Tolerability of NPA

Tolerability scores were obtained from nurses for 541 children, from parents/caregivers for 537 children, and from 137 children. Nurses estimated the lowest absolute baseline pain assessment scores for children before the NPA procedure (median 1 out of 10 [0,3]), and parents/caregivers estimated the highest score (median 3 [1,6]) (**Table 2**; **Figure 2**). The median difference in discomfort/distress/pain scores between before and during the NPA procedure was 3 [0,4], 2 [0,4] and 4 [2,4] as assessed by nurses, parents/caregivers, and children, respectively (**Table 2**).

**Figure 2.**
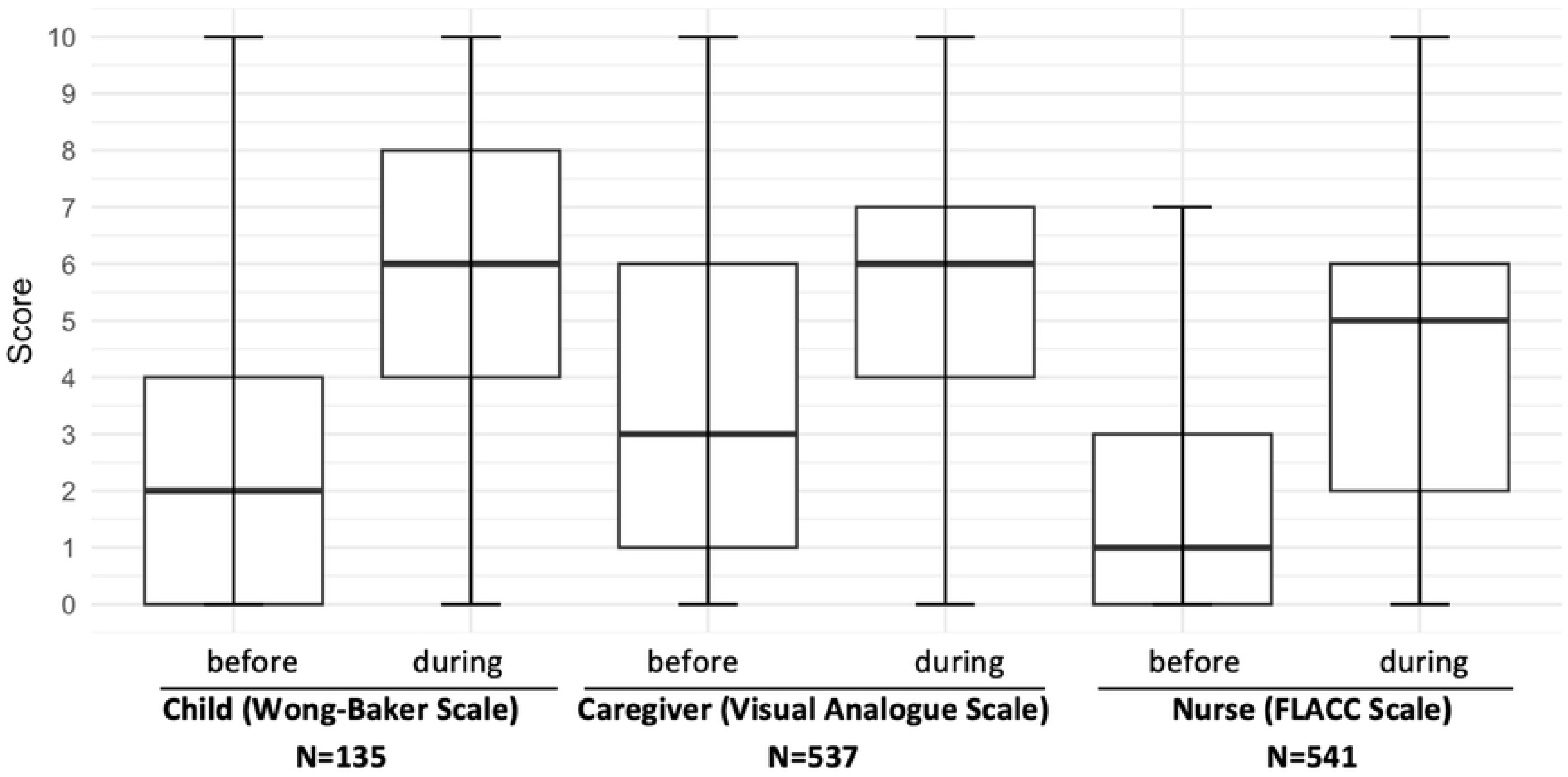
Absolute discomfort/distress/pain score before & during NPA collection assessed by children, parents/caregivers, nurses *FLACC: Face Legs Activity Cry Consolability scale; NPA: Nasopharyngeal aspiration; VAS: Visual Analog Scale: WBF: Wong Baker Face scale* *Data are min, Q1, median, Q3, max*

The median difference in nurse-assessed tolerability scores before and during the NPA sampling procedure varied largely between countries (p<0.001) and hospitals (p<0.001). Median differences of FLACC scores changed significantly according to the hospital’s experience in performing NPA, and the nurse’s assessment of NPA difficulty and child manageability. The was no difference according to the presence of blood in the NPA sample, the need for a repeat NPA in the second nostril, the presence of fast breathing and the child’s nutritional or HIV status (**Table 3**).

**Table 3.**
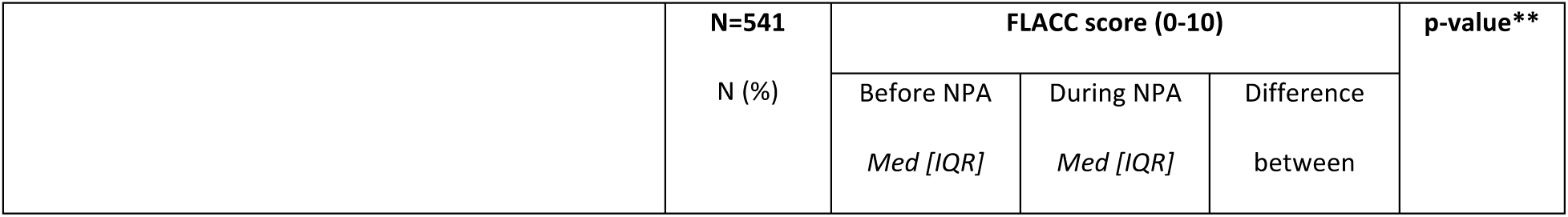
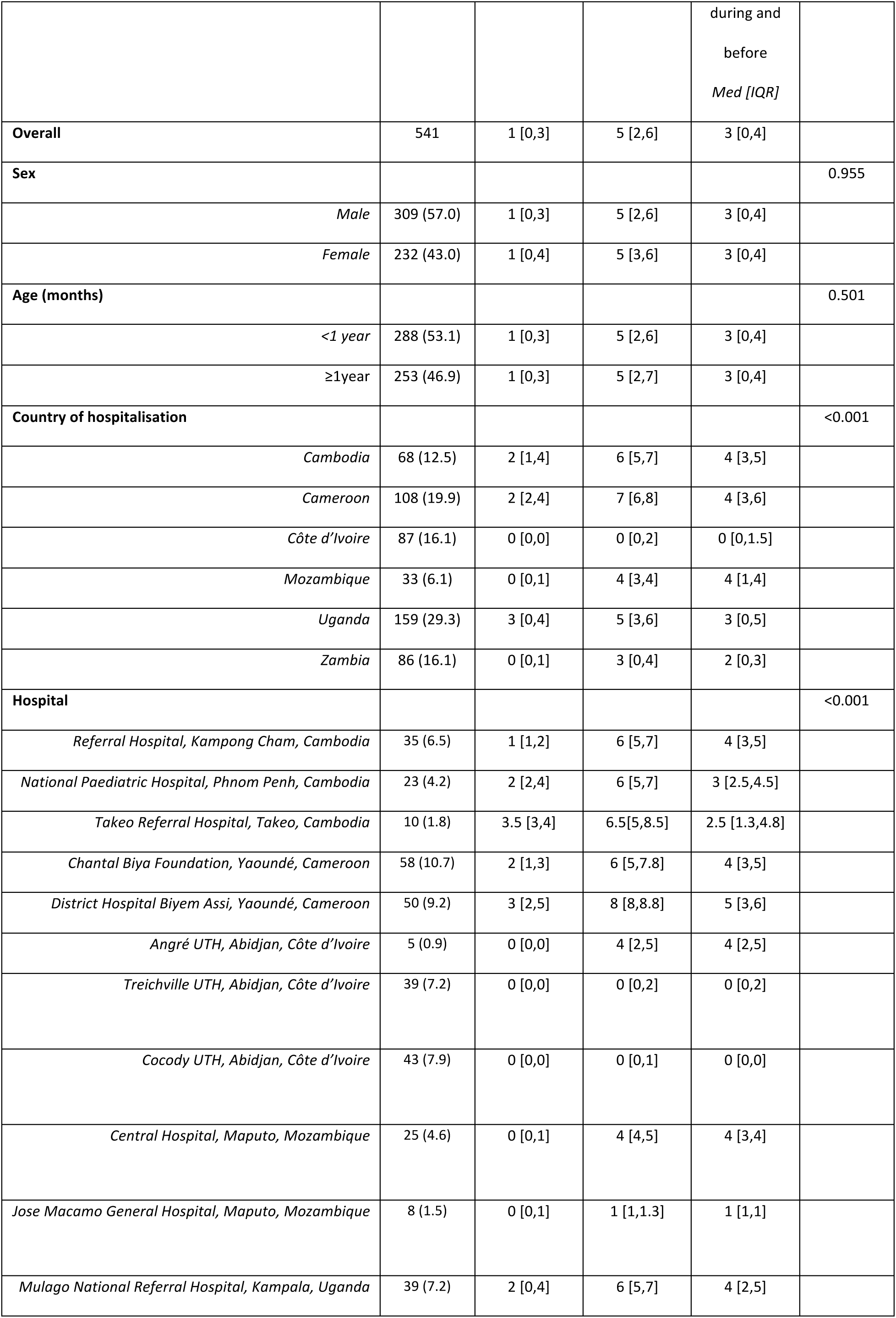
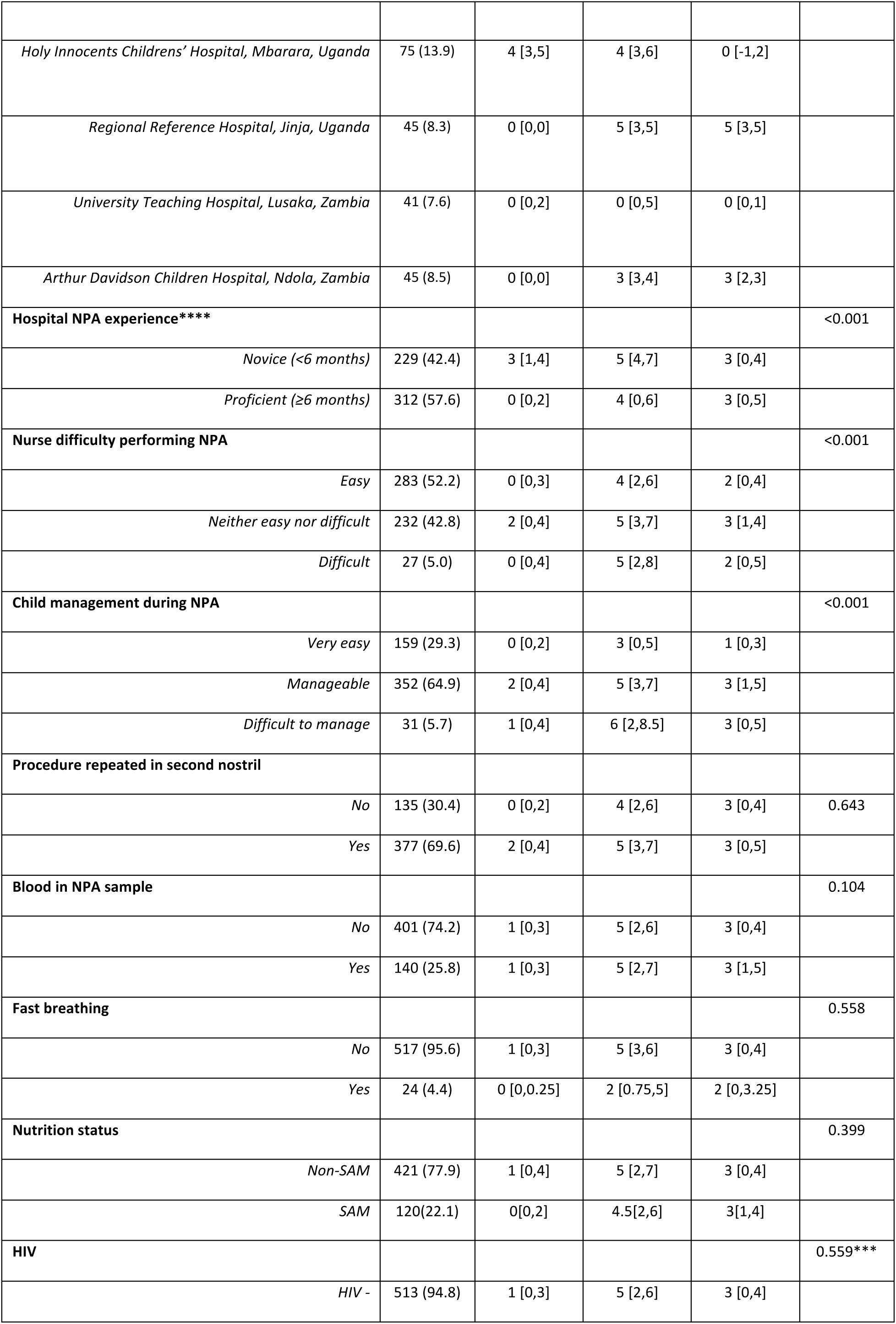
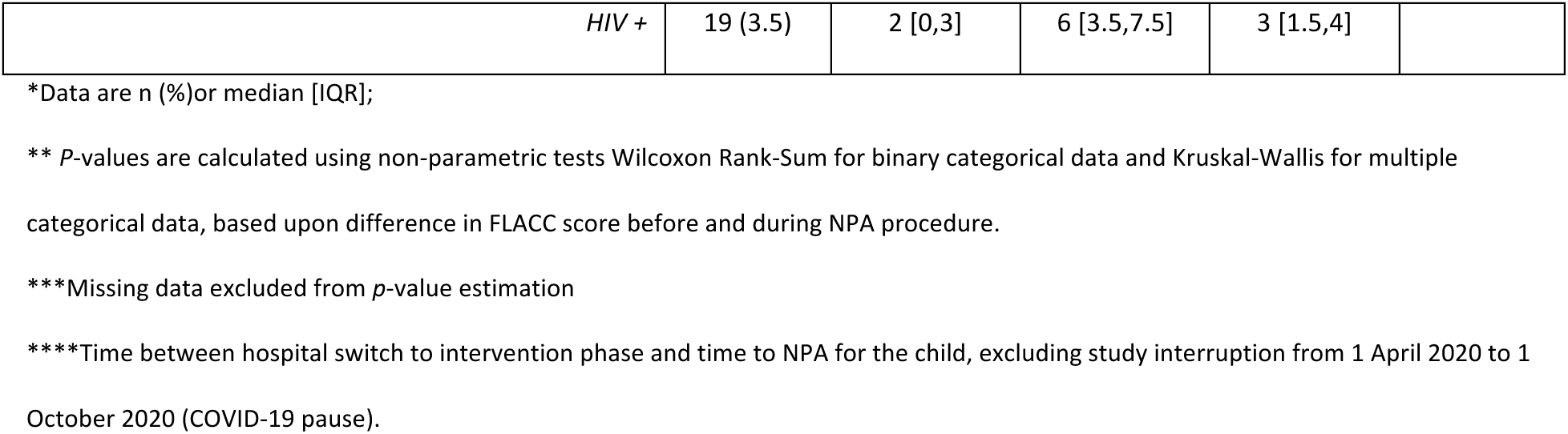
Discomfort/distress/pain score as assessed by nurse during versus before nasopharyngeal sampling procedure.

Four hospitals reported outlying FLACC assessment, consistently reported no change in or decrease in discomfort/distress/pain during the procedure compared to before. In these sites, children reported a median increase in pain 2 [2,4] during the procedure relative to before, but nurses and caregivers reported little or no difference, with median difference in score 0 [0,1] and 1 [-2,2] respectively (**Supplementary table 4 and 5**). Within the remaining 11 hospitals consistently reporting worse (increased) FLACC scores during the NPA procedure compared to before NPA, and which yielded normally distributed data (**Supplementary figure 2**), multivariable analysis showed a significant association between hospital and nurse-perceived difficulty in performing the procedure and poor tolerability (**Supplementary table 6**).

### Perceived feasibility and acceptability of NPA sample collection by nurses and parents/caregivers

Sixty-three nurses of median age 33.5 years (25-58 years) were interviewed, including 48 (76%) women; all were study nurses specifically recruited for TB-Speed Pneumonia, except 7 in Cameroon, who were hospital nurses. Their experience in the TB-Speed Pneumonia trial ranged from 1 month to 2 years. In addition, 59 parents/caregivers with a median age of 34.5 years (18–56) were interviewed, including 10 fathers (**Supplementary table 7).** No refusals to participate were reported.

#### • NPA procedure accepted but not always well understood

Parents/caregivers reported an overall positive acceptance of NPA sampling. They appreciated the novelty of the NPA procedure: “*…earlier I didn’t even know that this exam even exists, at the nose level, so I was really happy at least to be there for that (NPA) exam, I saw that it was really interesting” (Cote d’Ivoire, parent/caregiver)*. Parents/caregivers trusted nurses’ skills and intent during the NPA procedure*: “No, I trust them. He/she does not make my child in trouble, they treat my child to be cured” (Cambodia, parent/caregiver).* However, some parents/caregivers had limited understanding of the tuberculosis diagnostic purpose of NPA. NPA was seen as a procedure which mechanistically helped to reduce their child’s runny nose/nose blockage, and facilitated their breathing: “*Due to the reasons that before she had difficulty in breathing via throat and nose. After aspirating sputum, she can breathe as normal” (Cambodia, parent/caregiver).* Lack of awareness and stigma associated with tuberculosis among parents/caregivers was perceived by nurses as barriers to NPA acceptability: *“Many people, when you tell them to do the test, they say a difficult disease (tuberculosis) like that, a nasty disease (tuberculosis) like that, you shouldn’t put it on their children. You tell them it’s not necessarily that your children have this disease, but it’s to diagnose early, to see if your children have this disease” (Cote d’Ivoire, nurse)*.

#### • NPA experienced as unpleasant and sometimes painful, but benefitting child’s health

Many nurses and parents/caregivers talked about NPA as an unpleasant and even painful procedure for the child (and for themselves indirectly), that required repeated aspiration, could cause discomfort to the child and produce bloody samples: *“but you know when you inflict pain, it may not be pain but the discomfort, blood coming out, but those things happen, that is what I didn’t like” (Uganda, nurse,).* The key trigger allowing them to overcome this real/perceived pain for children was the perceived benefits for the child’s health: *“Uhh… even if the procedure is painful for some children, it does not harm anyone and we are doing it for the purpose of detecting tuberculosis and save lives on time…” (Cameroon, nurse)*.

#### • NPA sample collection perceived as feasible by nurses, with support from colleagues and parents/caregivers

In spite of discomfort caused to children, most nurses believed that, NPA was less invasive and quicker to perform than other sample collection procedures like gastric aspirate*: “The size of the probe that we use is also different…The gastric aspirate for example, is more invasive and takes a longer time, NPA is less painful for the child herself” (Mozambique, nurse).* Some nurses reported difficulties to collect sputum from very small children because of small nostrils; others found NPA easier to perform in younger children as compared to the older ones who resist and struggle during the procedure. In Cambodia in particular, nurses reported being frustrated with the protocol for NPA procedures, i.e., would have liked to use Vaseline and saline solution for NPA sample collection.

Almost all nurses reported the need for additional support from a colleague or parents/caregivers to restrain the child and/or to reassure the child during NPA procedure*: “…it is really necessary to immobilize them. Sometimes we often find ourselves calling a colleague or the parent himself, so we end up with two or three people only to immobilize a little boy” (Cameroon, nurse)*. Several parents/caregivers appreciated being asked for assistance by the nurses, and felt themselves engaged in the procedure: *“They said that please hold your child’s hands and feet tightly, to avoid movement. It would make it easy for doctor/nurse to collect NPA and I liked that, during NPA I wanted to stay with my child” (Cambodia, parent/caregiver)*.

#### • Education, continuous training and mentoring for NPA implementation

Parents/caregivers and nurses valued the importance of education, sharing information and reassurance throughout the NPA procedure: *“That was a new procedure in my life and I was sceptical in the beginning about having it performed on my child but after the information was given to me which I understood and I accepted to do for it (Zambia, parent/caregiver).* Nurses reported appreciating being trained on novel diagnostic tools, and improving their skills through day-to-day practice and experience: “*It was my first experience and training, so I gained a lot of skills during performing NPA, so I think I am equipped with enough skills and ideas about doing that procedure” (Uganda, nurse)*. Some nurses however regretted not being provided practical and refresher trainings to sustain their confidence and efficacy in obtaining adequate NPA samples: “*It (training) was I think just for a day, so I think we should have been trained more, we need to have just enough time and have enough practice on the patients (Zambia, nurse).* Nurses of all the countries valued the opportunity of collaboration and support from colleagues: in Cote d’Ivoire nurses shared practical tips, such as the practice of washing the nostril to moisten it and make NPA sample collection easier to perform.

### Perceived feasibility and acceptability of stool sample collection by nurses and parents/caregivers

#### • Stool sample collection as new non-invasive diagnostic sample for tuberculosis

Although stool testing was a well-known diagnostic test for childhood diseases, many parents/caregivers, and even nurses, were unaware that stool samples could be tested for tuberculosis: *“… there is something I don’t understand. Why did they ask for stool to diagnose tuberculosis? I know they have collected blood and there are some parameters they will check, they have done the NPA collection, and I am OK with that. But for the stool, I don’t really understand why they asked for it” (Cameroon, parent/caregiver)*.

The majority of parents/caregivers reported positive feelings and a good experience of stool sampling, as an easy and straightforward sample collection method: “*No, there was no complication there, (Laughs) there was nothing, they just gave me the box and I too gave what I had to give, that’s all, it was easy, I felt nothing” (Cameroon, parent/caregiver).* Nurses also appreciated the fact that no pain was caused to babies during stool sample collection (compared to NPA), and that no equipment was needed: *“Stool collection in children is a procedure which is simple, easily done it’s not painful both to the nurse and to the child …. (it) is easy and even cheap I think, every doctor every concerned person could make it a request to diagnose for tuberculosis especially in children” (Uganda, nurse).* Some nurses perceived that parents/caregivers would prefer stool collection to NPA, as they would feel more involved in the care provided: “*I think that if we present them all the methods, they will definitely choose on the stool sample. Yes, everyone will accept it very easily. Stools are easier for parents to accept compared to NPA, since the stool is collected by themselves” (Cameroon, nurse*). They also underlined that stool samples can be collected from any child (whether they are very sick, having difficulty in breathing or are under oxygen) as long as the child can produce stool.

#### • Frustrations due to time and conditions for sample collection

The perception of ease of collection was counterbalanced by the frustrations with delays in obtaining the stool samples*: “Some mothers can collect the stool and forget to tell you, they collect the sample in the night, and they tell you about it in the next day evening only” (Zambia, nurse)*. All respondents mentioned that children sometimes failed to produce stool during the hospitalization. Furthermore, both parents/caregivers and nurses experienced challenges with the collection procedure. The quantity of stool collected sometimes did not allow proper testing in the lab: “*For the stool I think it was not adequate because it is hard to know the quantity needed. Because it is when the mothers put in a little and the lab technicians say it was really very little. So, for measuring the quantity it is quite challenging” (Uganda, nurse); “It’s difficult to scrape stool from a diaper, if its watery and this causes problems to get adequate stool to send to the laboratory” (Mozambique, nurse).* Some nurses mentioned the risk of getting contaminated stool when it is collected by parents and not done appropriately: *“To me I would wish that nurses would have been the ones to collect stool, but these babies are always with mothers and they pass stool they do not use good clean techniques, they introduce other bacteria to that sample” (Uganda nurse).* Nurses recommended that laboratory staff, experienced in handling stool samples, would be well placed to provide continuous support to nurses and parents to improve the quality of sampling processes: “*I think they would call these lab people because they have more experience than us, if you continue training you adapt but more people could do it better would be the lab people because they know how they handle this sample and how to keep it so that it cannot get spoilt (Uganda, nurse)*.

## Discussion

This study indicated that collecting NPA and stool samples was highly feasible and well accepted by nurses and parents/caregivers for diagnosis of tuberculosis in young children hospitalized with severe pneumonia. Despite the vulnerability of the population, the safety of the NPA was good with very few and transient severe adverse events. NPA was frequently experienced as a moderately uncomfortable/painful procedure for children, which was counterbalanced by the perceptions of its benefits in terms of improving child health. The acceptability of stool as a non-invasive sample was high, however counterbalanced by low uptake, due to challenges with sample quality and delays in collection, and by slightly lower detection yield than NPA.

The lower uptake of stool sampling compared to NPA documented in our population of hospitalized children with severe pneumonia is in line with previous reports showing higher uptake of NPA than stool samples in children with presumptive tuberculosis at primary and secondary health care facilities, including mostly outpatient children in several high tuberculosis incidence countries (18, 19). Within our study, despite being admitted, many children were unable to pass stool during their time hospitalized, which was sometimes experienced as a frustration and a burden by nurses. The lower uptake of stool sampling compared to NPA needs to be balanced with the perceived invasiveness and cost of NPA (20).

Overall, the microbiological yield was low for both sample collection methods, in line with the expected low prevalence of tuberculosis in young children with severe pneumonia (21), but NPA and stool sampling and Ultra testing contributed to microbiological confirmation in more than 25% of children diagnosed with tuberculosis overall. Although stool had a lower yield than NPA, its incremental yield when combined to NPA support the recent WHO recommendation of rapid molecular testing on one stool and one respiratory sample in children (8).

Tolerability assessment of NPA among vulnerable children with severe pneumonia showed an increase, however modest (≤3 points out of 10), in discomfort/distress/pain scores in infants and young children during the NPA procedure, regardless of the assessor. Despite this increase in discomfort/distress/pain during the procedure, NPA was overall well tolerated by these vulnerable children with the exception of bloodstains in NPA samples, which are well-known minor adverse events in NPAs (22), and very few severe transient dyspnea. Nurses tended to report lower discomfort/distress/pain scores during the NPA procedure than parents/caregivers or children, which is the opposite of that reported by nurses from another NPA and stool study among children living with HIV without presumptive tuberculosis (23). In our study, nurses (unlike parents/caregivers) did not have knowledge of the child’s status and attitude when healthy, which would have informed their baseline assessment of pain, compared to pain being higher than normal during the procedure. It is also possible that nurses may have overestimated the tolerability of NPA in line with their perceived benefits of the procedure.

Contextual factors, namely the hospital and nurses’ experience in performing NPA, influenced the discomfort/distress/pain scores during NPA procedure. The increase in the nurse-assessed discomfort/distress/pain score during the NPA procedure was independently related to the hospital where the NPA was performed. Interestingly, nurses were more likely to report lower tolerability scores during the NPA procedure if they experienced difficulty performing the procedure. Difficulties met by nurses during sampling may have negatively impacted their ability to properly assess NPA tolerability. Studies have shown several gaps in HCWs capacity to assess and manage pain in children hospitalised in resource-limited settings (24, 25). Quality training and regular clinical mentoring is crucial for effective implementation of new clinical and technical interventions (26). Furthermore, child demographic and medical factors did not cause an important change in NPA tolerability as assessed by nurses. Relative to the range of the scores used (0–10), the median increase in pain across all scores during the procedure compared to baseline was modest (≤3 points). These findings suggests that with proper training of HCWs and proper preparation of parents/caregivers and children, NPA may be a good respiratory sample for diagnosis of tuberculosis in this vulnerable population.

Nurses reported positive attitudes towards NPA, perceiving it as less invasive than gastric aspirate and as contributing to rapid test results. Such positive attitudes at tertiary level of care match those reported in another TB-Speed study evaluating NPA at primary and secondary levels of care (12). Parents/caregivers did not understand that NPA was collected for tuberculosis diagnosis. One key driver of the acceptability of NPA nonetheless was the perception of positive health outcomes for the children; whether in terms of immediate improved breathing of children, as reported by both parents/caregivers and nurses, or in terms of capacity to diagnose or rule out tuberculosis, as reported by nurses (facilitating tuberculosis diagnosis thus improving the quality of care for these severely ill children). Despite a lack of understanding of the aim of stool sample collection procedure, parent’s acceptability of stool sample collection was good. In Ethiopia, caregivers interviewed on sample preferences for tuberculosis diagnosis reported preferring stool sampling compared to sputum, despite facing challenges in collecting stools on-the-spot (27). Overall, all parents/caregivers reported positive perceptions of nurse’s reassuring behaviour and advice/motivation during the management of their child, which probably contributed to the acceptability of the overall diagnostic package, with both stool and NPA sampling. In a study examining repeated tuberculosis sample collection in adults, an uncaring attitude and poor communication from HCWs acted as a barrier to the acceptability of tuberculosis investigations (28).

The study had several limitations. First, we did not assess time to NPA or stool sample collection within the feasibility assessment, thus are unable to compare the rapidity of both procedures in providing sample to be tested. Further the tolerability assessment of NPA based on pain scales, was conducted at 2 timepoints, “before/during”, only. Transience of the increase in discomfort/distress/pain was not assessed and we did not have any measure of “right after” pain. However, it is likely that increase in discomfort/distress/pain was transient, as it was reported in previous study. In addition, we conducted interviews in parents/caregivers who accepted these diagnostic procedures in the context of the trial, which may have biased reports toward more positive perceptions and experiences. Similarly, among HCWs, we only interviewed nurses, so we may have missed important perspectives from other key informants such as medical doctors or specialists. Furthermore, the study was conducted in tertiary hospitals with study nurses, thus not representing routine care conditions; the nurses’ reported experience of NPA and stool sample collection may therefore have been positively biased, as they were not likely to experience additional workload or burden related to the integration of new procedures within routine care.

Overall, our study provides important evidence on sample collection methods for microbiological tuberculosis diagnosis in children. Our data suggest that the NPA procedure is both safe and feasible in children with respiratory distress, which is highly promising considering that NPA can also be used for other microbiological tests beyond tuberculosis. The multi-country design of this sub-study supports the generalizability of the findings. The mixed methods approach contributes to internal validation of findings, with data saturation reached within the qualitative component, and a strong level of consistency between the quantitative and qualitative feasibility and tolerability findings. Although our in-depth documentation on NPA and stool is for an indication that is not yet validated (systematic detection of tuberculosis in children with severe pneumonia) and in a population that is not classically targeted for tuberculosis investigation, it does investigate the age group (< 5 years) for whom it is most challenging to obtain sputum, and for whom new sample types are most needed. Despite the WHO recommendation of rapid molecular testing on NPA among children (4), the rollout of NPA by national tuberculosis programs has globally been slower than that of stool testing (29). Although it would require more evidence on feasibility and acceptability in routine programmatic settings, our data indicating good feasibility, safety, and acceptability of NPA. Finally, our study provides additional evidence of the yield and feasibility of the Xpert Ultra on NPA and stool that supports the recent WHO recommendation of rapid molecular testing on one respiratory sample and one stool for diagnosis of tuberculosis in children.

## ACKNOWLEDGEMENTS

We would like to thank all the study nurses and parents/caregivers who provided their time to participate in this study. We would like to thank all the social science research assistants who conducted the data collection. We would also like to express our gratitude to Melanie Plazy from University of Bordeaux who contributed in developing tools of this study. Likewise, the authors would also like to express thanks to all TB-Speed staff who coordinated this acceptability and feasibility, many of whom are listed as part of the TB-Speed Pneumonia Study Group; the members of the TB-Speed Scientific Advisory Board who gave technical advice on the design of the study and approved the protocol: Stephen M Graham (University of Melbourne, Melbourne, VIC, Australia), Anneke Hesseling (Stellenbosch University, Cape Town, South Africa), Luis Cuevas (Liverpool School of Tropical Medicine, Liverpool, UK), Christophe Delacourt (Hopital Necker-Enfants Malades, Paris, France), Sabine Verkuijl (WHO, Geneva, Switzerland), Philippa Musoke (Makerere University, Kampala, Uganda), Mark Nicol (University of Western Australia, Perth, WA, Australia), Elizabeth Maleche-Obimbo (University of Nairobi, Nairobi, Kenya), and Chishala Chabala (University of Zambia, Lusaka, Zambia) who represented other TB-Speed investigators at Scientific Advisory Board meetings; the Ministries of Health and National Tuberculosis Programmes (NTPs) of participating countries; and the NTP district representatives who were supporting the TB-Speed Pneumonia Study implementation.

## DATA AVAILABILITY STATEMENT

Persons wishing to have access to the TB-Speed Pneumonia study data, including clinical data, questionnaires and interviews, may submit a request to the corresponding author, who will ensure the request is duly examined by the TB-Speed publication committee and the study sponsor.

## REFERENCES

1. World Health Organisation. Global tuberculosis report 2024. Geneva: World Health Organization; 2024. Available from: https://iris.who.int/server/api/core/bitstreams/7292c91e-ffb0-4cef-ac39-0200f06961ea/content.

2. Dodd PJ, Yuen CM, Sismanidis C, Seddon JA, Jenkins HE. The global burden of tuberculosis mortality in children: a mathematical modelling study. Lancet Glob Health. 2017;5(9):e898–e906. doi: 10.1016/S2214-109X(17)30289-9

3. Wobudeya E, Bonnet M, Walters EG, Nabeta P, Song R, Murithi W, et al. Diagnostic Advances in Childhood Tuberculosis-Improving Specimen Collection and Yield of Microbiological Diagnosis for Intrathoracic Tuberculosis. Pathogens. 2022;11(4). doi: 10.3390/pathogens11040389

4. World Health Organisation. WHO consolidated guidelines on tuberculosis. Module 5: management of tuberculosis in children and adolescents. Geneva: World Health Organization; 2022. Available from: https://iris.who.int/server/api/core/bitstreams/6212d328-f45e-4905-ae32-e0161d4f0029/content.

5. Moore BK, Graham SM, Nandakumar S, Doyle J, Maloney SA. Pediatric Tuberculosis: A Review of Evidence-Based Best Practices for Clinicians and Health Care Providers. Pathogens. 2024;13(6). doi: 10.3390/pathogens13060467

6. Khambati N, Song R, MacLean EL, Kohli M, Olbrich L, Bijker EM. The diagnostic yield of nasopharyngeal aspirate for pediatric pulmonary tuberculosis: a systematic review and meta-analysis. BMC Glob Public Health. 2023;1. doi: 10.1186/s44263-023-00018-1

7. Olbrich L, Yang B, Poore H, Razid A, Sweetser B, Damkjaer MW, et al. Parallel use of low-complexity automated nucleic acid amplification tests on respiratory and stool samples with or without lateral flow lipoarabinomannan assays to detect pulmonary tuberculosis disease in children. Cochrane Database Syst Rev. 2025;6(6):CD016071. doi: 10.1002/14651858.CD016071.pub2

8. World Health Organisation. WHO consolidated guidelines on tuberculosis. Module 3: diagnosis. Geneva: World Health Organization; 2025. Available from: https://iris.who.int/server/api/core/bitstreams/ae20e43e-17fd-4951-b475-3e36e166e7ae/content.

9. Oliwa JN, Karumbi JM, Marais BJ, Madhi SA, Graham SM. Tuberculosis as a cause or comorbidity of childhood pneumonia in tuberculosis-endemic areas: a systematic review. Lancet Respir Med. 2015;3(3):235–43. doi: 10.1016/S2213-2600(15)00028-4

10. Nantongo JM, Wobudeya E, Mupere E, Joloba M, Ssengooba W, Kisembo HN, et al. High incidence of pulmonary tuberculosis in children admitted with severe pneumonia in Uganda. BMC Pediatr. 2013;13:16. doi: 10.1186/1471-2431-13-16

11. Marcy O, Wobudeya E, Font H, Vessiere A, Chabala C, Khosa C, et al. Effect of systematic tuberculosis detection on mortality in young children with severe pneumonia in countries with high incidence of tuberculosis: a stepped-wedge cluster-randomised trial. Lancet Infect Dis. 2023;23(3):341–51. doi: 10.1016/S1473-3099(22)00668-5

12. Joshi B, De Lima YV, Massom DM, Kaing S, Banga MF, Kamara ET, et al. Acceptability of decentralizing childhood tuberculosis diagnosis in low-income countries with high tuberculosis incidence: Experiences and perceptions from health care workers in Sub-Saharan Africa and South-East Asia. PLOS Glob Public Health. 2023;3(10):e0001525. doi: 10.1371/journal.pgph.0001525

13. Vessiere A, Font H, Gabillard D, Adonis-Koffi L, Borand L, Chabala C, et al. Impact of systematic early tuberculosis detection using Xpert MTB/RIF Ultra in children with severe pneumonia in high tuberculosis burden countries (TB-Speed pneumonia): a stepped wedge cluster randomized trial. BMC Pediatr. 2021;21(1):136. doi: 10.1186/s12887-021-02576-5

14. Division of AIDS, National Institute of Allergy and Infectious Diseases. Division of AIDS (DAIDS) Table for Grading the Severity of Adult and Pediatric Adverse Events. Corrected Version 2.1. July 2017. National Institutes of Health, US Department of Health and Human Services; 2017. Available from: https://rsc.niaid.nih.gov/sites/default/files/daidsgradingcorrectedv21.pdf.

15. Merkel SI, Voepel-Lewis T, Shayevitz JR, Malviya S. The FLACC: a behavioral scale for scoring postoperative pain in young children. Pediatr Nurs. 1997;23(3):293–7.

16. Garra G, Singer AJ, Domingo A, Thode HC, Jr. The Wong-Baker pain FACES scale measures pain, not fear. Pediatr Emerg Care. 2013;29(1):17–20. doi: 10.1097/PEC.0b013e31827b2299

17. Sekhon M, Cartwright M, Francis JJ. Acceptability of healthcare interventions: an overview of reviews and development of a theoretical framework. BMC Health Serv Res. 2017;17(1):88. doi: 10.1186/s12913-017-2031-8

18. Marcy O, Ung V, Goyet S, Borand L, Msellati P, Tejiokem M, et al. Performance of Xpert MTB/RIF and Alternative Specimen Collection Methods for the Diagnosis of Tuberculosis in HIV-Infected Children. Clin Infect Dis. 2016;62(9):1161–8. doi: 10.1093/cid/ciw036

19. Wobudeya E, Nanfuka M, Ton Nu Nguyet MH, Taguebue JV, Moh R, Breton G, et al. Effect of decentralising childhood tuberculosis diagnosis to primary health centre versus district hospital levels on disease detection in children from six high tuberculosis incidence countries: an operational research, pre-post intervention study. EClinicalMedicine. 2024;70:102527. doi: 10.1016/j.eclinm.2024.102527

20. d’Elbee M, Harker M, Mafirakureva N, Nanfuka M, Huyen Ton Nu Nguyet M, Taguebue JV, et al. Cost-effectiveness and budget impact of decentralising childhood tuberculosis diagnosis in six high tuberculosis incidence countries: a mathematical modelling study. EClinicalMedicine. 2024;70:102528. doi: 10.1016/j.eclinm.2024.102528

21. Pneumonia Etiology Research for Child Health Study G. Causes of severe pneumonia requiring hospital admission in children without HIV infection from Africa and Asia: the PERCH multi-country case-control study. Lancet. 2019;394(10200):757–79. doi: 10.1016/S0140-6736(19)30721-4

22. Owens S, Abdel-Rahman IE, Balyejusa S, Musoke P, Cooke RP, Parry CM, et al. Nasopharyngeal aspiration for diagnosis of pulmonary tuberculosis. Arch Dis Child. 2007;92(8):693–6. doi: 10.1136/adc.2006.108308

23. Sanogo B, Kiema PE, Barro M, Nacro SF, Ouermi SA, Msellati P, et al. Contribution and Acceptability of Bacteriological Collection Tools in the Diagnosis of Tuberculosis in Children Infected with HIV. J Trop Pediatr. 2021;67(2). doi: 10.1093/tropej/fmab027

24. Wuni A, Salia SM, Mohammed Ibrahim M, Iddriss I, Abena Nyarko B, Nabila Seini S, et al. Evaluating Knowledge, Practices, and Barriers of Paediatric Pain Management among Nurses in a Tertiary Health Facility in the Northern Region of Ghana: A Descriptive Cross-Sectional Study. Pain Res Manag. 2020;2020:8846599. doi: 10.1155/2020/8846599

25. Kusi Amponsah A, Hammond CK, Bam V, Gyamfi D, Armah J, Wilson D, et al. Implementation and Evaluation of a Pediatric Pain Assessment Educational Program (PPAEP) for Nurses in a Resource-Limited Setting: A Pilot Study. Paediatr Neonatal Pain. 2026;8(2):e70027. doi: 10.1002/pne2.70027

26. Feyissa GT, Balabanova D, Woldie M. How Effective are Mentoring Programs for Improving Health Worker Competence and Institutional Performance in Africa? A Systematic Review of Quantitative Evidence. J Multidiscip Healthc. 2019;12:989–1005. doi: 10.2147/JMDH.S228951

27. Yenew B, de Haas P, Babo Y, Diriba G, Sherefdin B, Bedru A, et al. Diagnostic accuracy, feasibility and acceptability of stool-based testing for childhood tuberculosis. ERJ Open Res. 2024;10(3). doi: 10.1183/23120541.00710-2023

28. Kumwenda M, Nyang’wa BT, Chikuse B, Biseck T, Maosa S, Chilembwe A, et al. The second sputum sample complicates tuberculosis diagnosis for women: a qualitative study from Malawi. Int J Tuberc Lung Dis. 2017;21(12):1258–63. doi: 10.5588/ijtld.17.0146

29. Klinkenberg E, de Haas P, Manyonge C, Namutebi J, Mujangi B, Mutunzi H, et al. Lessons Learned from Early Implementation and Scale-up of Stool-Based Xpert Testing to Diagnose Tuberculosis in Children. Emerg Infect Dis. 2025;31(3):1–9. doi: 10.3201/eid3103.241580

